# Environmental Contamination of SARS-CoV-2 in a Non-Healthcare Setting Revealed by Sensitive Nested RT-PCR

**DOI:** 10.1101/2020.05.31.20107862

**Authors:** Judith Chui Ching Wong, Hapuarachchige Chanditha Hapuarachchi, Sathish Arivalan, Wei Ping Tien, Carmen Koo, Diyar Mailepessov, Marcella Kong, Mohammad Nazeem, Merrill Lim, Lee Ching Ng

**Affiliations:** National Environmental Agency, Environmental Health Institute, Singapore; School of Biological Sciences, Nanyang Technological University, Singapore

## Abstract

Fomite-mediated transmission has been identified as a possible route for disease spread of the COVID-19 pandemic. In healthcare settings, evidence of environmental contamination by SARS-CoV-2 has been found in patients’ rooms and toilets. Here, we investigate environmental contamination of SARS-CoV-2 in non-healthcare settings and assessed the efficacy of cleaning and disinfection in removing SARS-CoV-2 contamination. A total of 428 environmental swabs and six air samples was taken from accommodation rooms, toilets and elevators that have been used by COVID-19 cases. Through the use of a sensitive nested RT-PCR assay, we found two SARS-CoV-2 RNA positive samples from the room resided by a COVID-19 case, highlighting the risk of fomite-mediated transmission in non-healthcare settings and the importance of surface disinfection of spaces occupied by cases. Of note, we did not find evidence for air-borne transmission, nor of environmental contamination of elevators, which were transiently exposed to infected persons.

## Introduction

The coronavirus disease 2019 (COVID-19) first emerged in December 2019 in the city of Wuhan in Hubei Province, China, and has since spread to more than 200 countries across the globe (1). Recognised as a global pandemic by the World Health Organization (WHO) in March 2020 (2), the outbreak has caused over 3.2 million infections and 230,000 deaths to date. In Singapore, more than 16,000 COVID-19 cases have been reported as of April 2020, with local clusters of transmission occurring in a tourist shop, places of worship, business meetings, work places and worker dormitories, among others (3). COVID-19 is caused by the Severe Acute Respiratory Syndrome Coronavirus 2 (SARS-CoV-2) and common symptoms include fever, cough, fatigue and sputum production (4). Presently, there is no effective vaccine or treatment available to treat SARS-CoV-2 infections. Practicing good personal and environmental hygiene and social distancing are used as preventive measures.

Similar to other human coronaviruses, SARS-CoV-2 is thought to spread through person-to-person contact, respiratory droplets or through contact with contaminated surfaces (5). Evidence of environmental contamination has been shown in healthcare settings, where SARS-CoV (6, 7), Middle East Respiratory Syndrome Coronavirus (MERS-CoV) (8, 9), and more recently, SARS-CoV-2 (10), have been found around the patients’ bed and toilet areas. Laboratory tests also demonstrate that SARS-CoV-2 can survive on environmental surfaces for two to three days (11, 12), further emphasizing the risk of fomite-mediated transmission, particularly in the healthcare environments.

As the contaminated environment presents a possible route of transmission, the National Environment Agency introduced measures to reduce the risk of fomite-mediated transmission after the onset of the first few cases in Singapore. Within one week of the diagnosis of the first imported cases, the agency issued specific guidelines for cleaning and disinfection of premises and residences exposed to SARS-CoV-2 infected persons (13, 14), as well as advisories to increase frequency of general cleaning of premises. Supervision for cleaning and disinfection procedures was also provided for non-healthcare premises.

To assess the extent of surface contamination and the efficacy of disinfection procedures, environmental surface swabs were taken before and after cleaning and disinfection of common high-touchpoint areas in accommodation rooms, toilets and elevators of premises where infected persons resided or visited. As some reports suggest the possibility of aerosol-borne transmission (12, 15), air samples were also taken from an enclosed air-conditioned room that had no mechanical ventilation. We detected two SARS-CoV-2 RNA positive samples from the accommodation room used by a SARS-CoV-2 infected person prior to diagnosis, highlighting the risk of fomite-mediated transmission in non-healthcare settings.

## Results and Discussion

From 28 February 2020 to 20 March 2020, 428 environmental surface swabs were collected from 18 sites, comprising of four accommodation rooms (n=134) and eight toilets (n=120) and six elevators (n=174). The accommodation rooms and toilets were from a home residence (2 rooms, 1 toilet) and two commercial boarding residences (1 room, 1 toilet, and 1 room, 6 toilets, respectively), while the elevators were from public housing blocks (3 elevators per block). Persons confirmed to be infected with SARS-CoV-2 have resided or visited these sites prior to diagnosis and isolation (Table 1). As diagnosis and isolation are typically a few days after the onset of symptoms, it is assumed that the cases were symptomatic while residing or visiting these sites. Surfaces potentially exposed to the touch and any cough of an infected person (e.g. walls, table surfaces) were sampled. Air samples were collected (n=4) in an accommodation room (occupied by Case 1) that was thought to be poorly ventilated and another 2 samples were collected right outside the room entrance. All samples were taken after the infected persons vacated the sites and have been isolated in healthcare facilities. Half of the surface swab and air samples were taken before the cleaning and disinfection and the other half was taken after the disinfection procedure.

**Table 1.** Sampling premises, air-ventilation and disinfectants used.

| Case | No. of sites | No. of Samples<sup>1</sup> | Days after occupancy | Estimated floor area (m<sup>2</sup>) | Ventilation | Air-conditioning system | Active Ingredient in Disinfectants |
| --- | --- | --- | --- | --- | --- | --- | --- |
| 1<sup>2</sup> | Commercial Boarding Room (n=1)<br>Toilets (n=6) | 120<sup>3</sup> | 1 | Room - 4<br>Toilet – 1.2 | Mechanical ventilation limited to toilets only | Wall-mounted fan coil unit | Sodium chlorite |
| 2<sup>4</sup> | Home Residence Room (n=2)<br>Toilet (n=1) | 100 | 3 | Room 1– 12<br>Room 2 – 14.5<br>Toilet – 3.8 | Naturally ventilated, ambient temperature | Wall-mounted fan coil unit (not used since case vacated) | Hydrogen peroxide and peroxyacetic acid |
| 3<sup>4</sup> |  |  | 1 |  |  |  |  |
| 4 | Commercial Boarding Room (n=1)<br>Toilet (n=1) | 40 | 3 | Room – 50<br>Toilet - 5 | Mechanical ventilation | Fan coil unit | Benzalkonium chloride |
| 5 | Elevators of public housing (n=3) | 96 | 2 | 1.7 | Mechanical and natural, ambient temperature | Nil | Sodium hypochlorite |
| 6 | Elevators of public housing (n=3) | 78 | 3 | 1.7 | Mechanical and natural, ambient temperature | Nil | Sodium hypochlorite |

<sup>1</sup> Half of the surface swab and air samples were taken before the cleaning and disinfection and the other half was taken after the disinfection procedure, performed by third party commercial companies
<sup>2</sup> Case 1 resided in a residence with shared toilets
<sup>3</sup> Comprises of 114 swab samples and 6 air samples
<sup>4</sup> Cases 2 and 3 used two different rooms in a shared residence

Among 428 swab samples taken (Table 1), only two were positive for SARS-CoV-2 RNA (Table 2). They were from the bedside wall and bed handle of the accommodation room of Case 1, before disinfection and cleaning was carried out. Sequencing of the nested PCR fragments (356 bp) confirmed the environmental contamination of the room. The two positive samples yielded identical genetic sequence in the 356 bp fragment (Figure 1). The room was cleaned on the same day, and samples collected after the cleaning had undetectable SARS-CoV-2, suggesting the efficacy of the cleaning methodology. The presence of virus contamination on surfaces of a primary space occupied by a case for a prolonged period of time highlights the risk of fomite-mediated transmission and necessitates the regular cleaning and disinfection of environmental surfaces, particularly when COVID-19 transmission is present. The risk has been borne out by a report on the epidemiological link between two sets of cases who had not met but sat in the same seats in sequential services of a church in Singapore (16). The risk of environmental contamination is corroborated by studies in healthcare settings including a recent one on SARS-CoV-2 in a Singapore hospital where 60.7% of the samples were found to be positive, and previously on MERS-CoV (20.4%) and SARS-CoV (7.6%) (6, 8, 10). The higher contamination rate in hospitals can be attributed to the duration of stay of cases, and the viral shedding capacity of a patient at different stages of the disease.

**Table 2.**
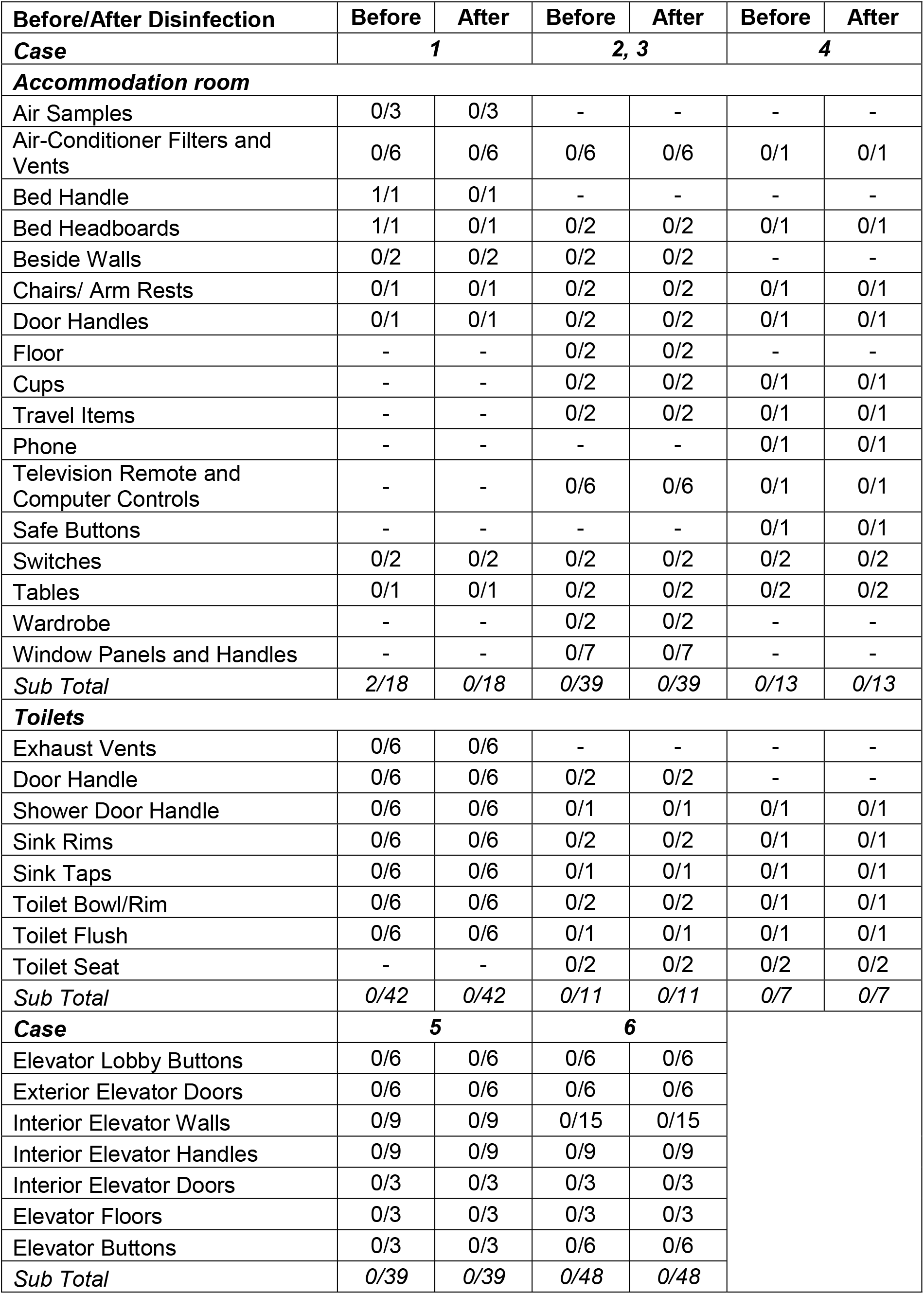
Environmental samples and corresponding nested RT-PCR results.

| Before/After Disinfection | Before | After | Before | After | Before | After |
| --- | --- | --- | --- | --- | --- | --- |
| Case | 1 |  | 2, 3 |  | 4 |  |
| Accommodation room |  |  |  |  |  |  |
| Air Samples | 0/3 | 0/3 | - | - | - | - |
| Air-Conditioner Filters and Vents | 0/6 | 0/6 | 0/6 | 0/6 | 0/1 | 0/1 |
| Bed Handle | 1/1 | 0/1 | - | - | - | - |
| Bed Headboards | 1/1 | 0/1 | 0/2 | 0/2 | 0/1 | 0/1 |
| Beside Walls | 0/2 | 0/2 | 0/2 | 0/2 | - | - |
| Chairs/ Arm Rests | 0/1 | 0/1 | 0/2 | 0/2 | 0/1 | 0/1 |
| Door Handles | 0/1 | 0/1 | 0/2 | 0/2 | 0/1 | 0/1 |
| Floor | - | - | 0/2 | 0/2 | - | - |
| Cups | - | - | 0/2 | 0/2 | 0/1 | 0/1 |
| Travel Items | - | - | 0/2 | 0/2 | 0/1 | 0/1 |
| Phone | - | - | - | - | 0/1 | 0/1 |
| Television Remote and Computer Controls | - | - | 0/6 | 0/6 | 0/1 | 0/1 |
| Safe Buttons | - | - | - | - | 0/1 | 0/1 |
| Switches | 0/2 | 0/2 | 0/2 | 0/2 | 0/2 | 0/2 |
| Tables | 0/1 | 0/1 | 0/2 | 0/2 | 0/2 | 0/2 |
| Wardrobe | - | - | 0/2 | 0/2 | - | - |
| Window Panels and Handles | - | - | 0/7 | 0/7 | - | - |
| Sub Total | 2/18 | 0/18 | 0/39 | 0/39 | 0/13 | 0/13 |
| Toilets |  |  |  |  |  |  |
| Exhaust Vents | 0/6 | 0/6 | - | - | - | - |
| Door Handle | 0/6 | 0/6 | 0/2 | 0/2 | - | - |
| Shower Door Handle | 0/6 | 0/6 | 0/1 | 0/1 | 0/1 | 0/1 |
| Sink Rims | 0/6 | 0/6 | 0/2 | 0/2 | 0/1 | 0/1 |
| Sink Taps | 0/6 | 0/6 | 0/1 | 0/1 | 0/1 | 0/1 |
| Toilet Bowl/Rim | 0/6 | 0/6 | 0/2 | 0/2 | 0/1 | 0/1 |
| Toilet Flush | 0/6 | 0/6 | 0/1 | 0/1 | 0/1 | 0/1 |
| Toilet Seat | - | - | 0/2 | 0/2 | 0/2 | 0/2 |
| Sub Total | 0/42 | 0/42 | 0/11 | 0/11 | 0/7 | 0/7 |
| Case | 5 |  | 6 |  |  |  |
| Elevator Lobby Buttons | 0/6 | 0/6 | 0/6 | 0/6 |  |  |
| Exterior Elevator Doors | 0/6 | 0/6 | 0/6 | 0/6 |  |  |
| Interior Elevator Walls | 0/9 | 0/9 | 0/15 | 0/15 |  |  |
| Interior Elevator Handles | 0/9 | 0/9 | 0/9 | 0/9 |  |  |
| Interior Elevator Doors | 0/3 | 0/3 | 0/3 | 0/3 |  |  |
| Elevator Floors | 0/3 | 0/3 | 0/3 | 0/3 |  |  |
| Elevator Buttons | 0/3 | 0/3 | 0/6 | 0/6 |  |  |
| Sub Total | 0/39 | 0/39 | 0/48 | 0/48 |  |  |

**Figure 1.**
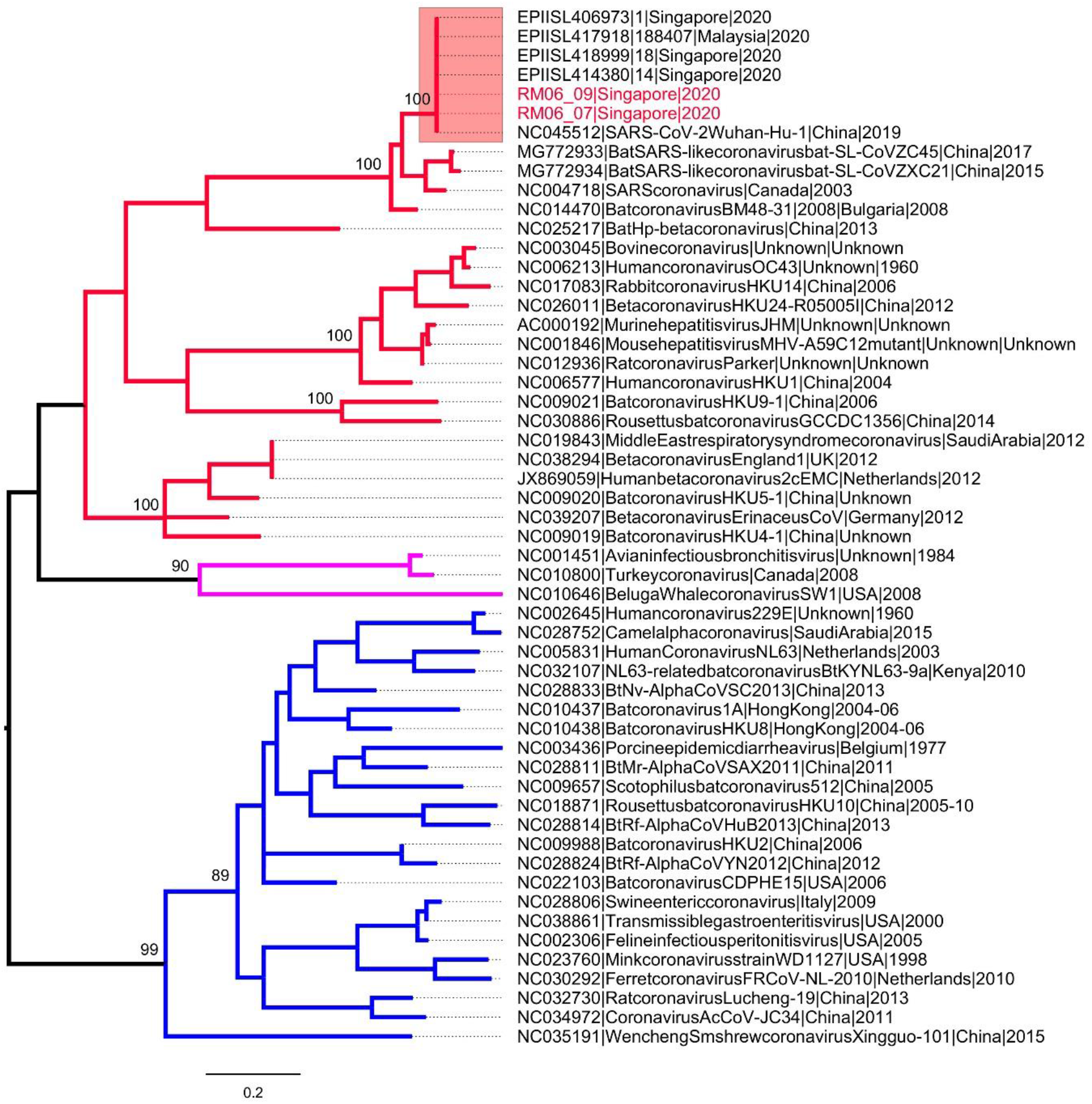
Phylogenetic analysis of RNA dependent RNA polymerase gene (356 bp) of coronaviruses. The maximum likelihood tree was constructed by using the general time reversible substitution model with gamma and invariant site parameters (GTR + Gamma 5 + I). Different groups of coronaviruses are shown in different branch colours; group 1 (blue), group 2 (red) and group 3 (purple). SARS-CoV-2 sequences are highlighted in the pink box. The taxa names of SARS-CoV-2 sequences obtained from two positive swabs are highlighted in red. The numbers shown on nodes are bootstrap values.

SARS-CoV-2 was not detected in all other pre- and post-cleaning samples (n=426). The general low positive rate, even before cleaning and disinfection, demonstrates the challenge in environmental sampling and detection, especially due to the time lapse between exposure and sampling. The positive accommodation room was the only site where sampling was performed a day after a relatively long period of exposure to a case. The small area (4 m^2^) of the affected room probably increased the probability of prolonged contamination and positive sampling. Although sampling for Case 1 was performed only one day after the case vacated the room, the case spent a day with symptoms in the room.

The negative results in common areas, such as elevators and their call buttons (for Cases 5-6) as well as shared toilets (for Cases 1-3), even before cleaning could possibly be contributed by the following factors: i) cases had not dwelled in these areas for a prolonged period of time, and the transient exposure did not lead to a severe contamination; ii) the frequent cleaning regime implemented since Singapore received imported cases had mitigated the risk; iii) the warm and humid tropical climate has reduced the contamination, especially in naturally ventilated spaces. These are perhaps the factors that have led to the contrasting results from the 2003 finding at the Hong Kong Metropole hotel elevator area, where SARS-CoV viral RNA was detected even three months after the case had left the hotel (17).

Laboratory tests have shown that SARS-CoV-2 could be found in aerosols for only 2-3 hours (12). In our study, expectedly, SARS-CoV-2 was not detected in any of the air samples, since premises were vacated at least 24 hours prior to sampling. On a similar note, SARS-CoV-2 was not detected in the return or supply air vents of different air-conditioner systems, in contrast to a previous case report in a hospital isolation room (10). In the latter case report, high viral shedding from the resident patient, air flow pattern and exhaust outlet location were thought to be possible factors for the virus to be found on the exhaust outlet. Further investigations in non-health care settings is needed for risk assessment.

As environmental samples often contain low amounts of viral RNA, we developed a sensitive nested PCR protocol for the detection of SARS-CoV-2 RNA, modified from primers published by Woo *et. al*. for generic coronavirus. Using synthetic RNA template with known copy numbers (1+E6 copies per μL; Twist Bioscience, USA), we show that it has a sensitivity of 12 RNA copies in a sample, equivalent to the China CDC assay and more sensitive than the HKU and US CDC protocols (Supplementary Table 1). Besides being highly sensitive, the nested protocol generated genomic fragments long enough to be sequenced. We also tested the two positive samples by using the two probe-based real-time PCR assays developed by Hong Kong University (HKU) and recommended by the World Health Organization for the detection of SARS-CoV-2 (18). Both assays, nucleocapsid protein (screening assay; 110 bp) and open reading frame 1b (confirmatory assay; 132 bp), gave positive results for the two environmental samples and indicated approximately 400 RNA copy numbers in each sample.

To our knowledge, this is the first study of environmental contamination in non-healthcare premises. The detection of viral RNA in the accommodation room of a confirmed case highlights the importance of disinfection in the community, especially primary spaces of cases, to avoid fomite-mediated transmission. While the study did not reveal gross contamination of the environment, especially secondary sites visited by the cases, high frequency general cleaning of common areas has and will likely mitigate transmission in the community.

## Materials and Methods

### Environmental samples

Environmental surface swabs were taken from common high-touchpoint areas in accommodation rooms, toilets and elevators of premises (Table 1, Figure 2). Pre-moistened sterile synthetic-tipped swabs were used for the surface sampling. The swabs were placed in tubes containing 500 μL Viral Transport Media (VTM). Field blanks were taken by moistening the swab and placing into the tubes with VTM, without any surface sampling. These were done at each sampling site to determine if any cross-contamination has occurred during sample collection. Details of the sampling premises as well as the active ingredients of the disinfectants used for surface disinfection are listed in Table 1.

**Figure 2.**
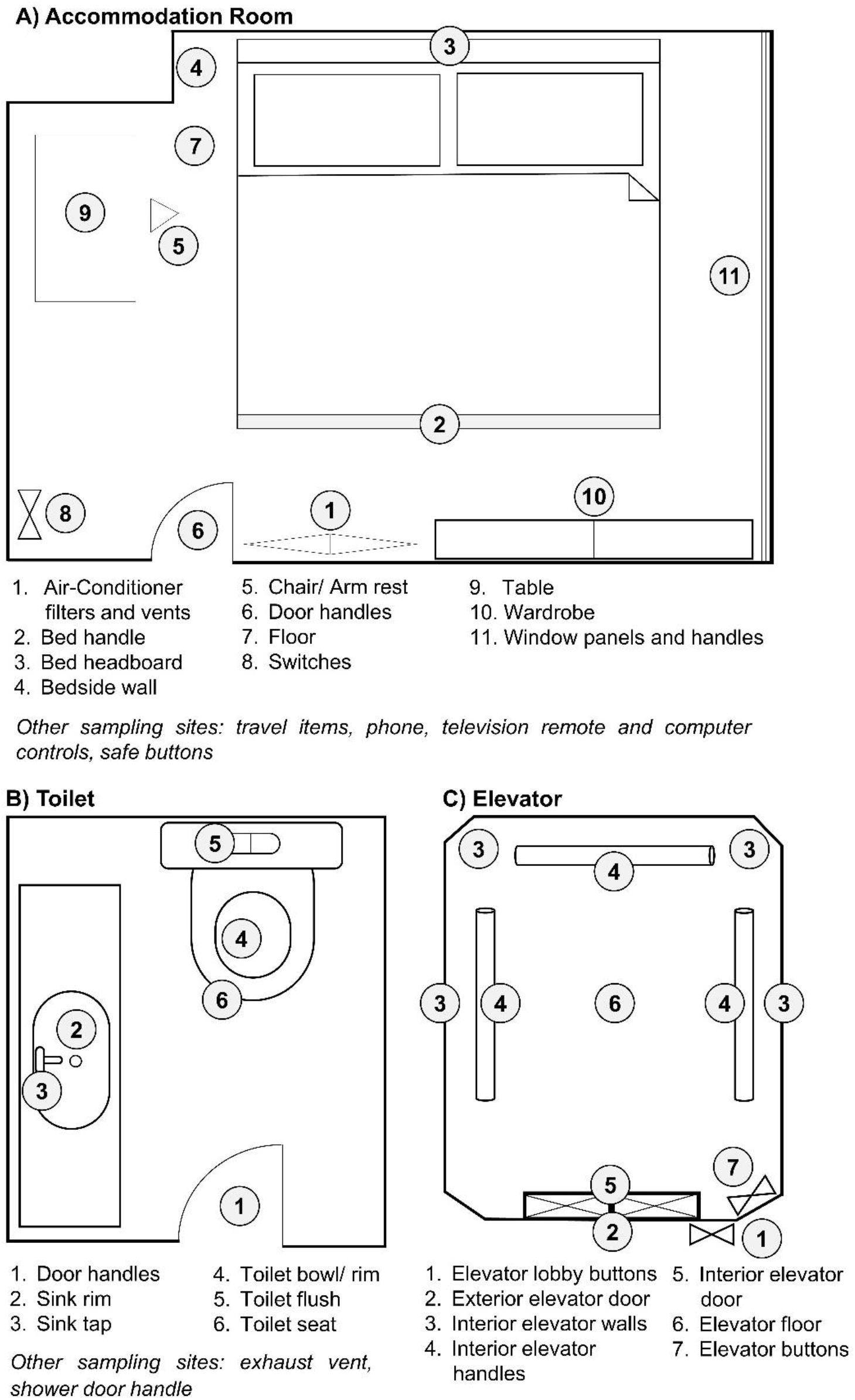
Representative images of sampling premises. Representative sampling sites for A – Accommodation Rooms, B – Toilets and C – Elevators. Environmental swab samples were taken at the areas indicated.

Air samples were collected in VTM using a cyclonic air sampler (Coriolis, Bertin Instruments) with an air flow rate of 300L/min for a duration of 30 mins per sample.

### Extraction of SARS-CoV-2 RNA from environmental samples

Swabs in VTM were vortexed at high speed for 60 secs (19). The wet swabs were pressed against the innerwall of the tube to squeeze out as much liquid as possible. The remaining solution was centrifuged at 13,000 rpm for 1 min to remove any debris. A 140 μL aliquot of the supernatant was used for the extraction of viral RNA by using the QIAMP Viral RNA Mini Kit (Qiagen, Hilden, Germany) according to the protocol recommended by the manufacturer. For air samples collected in VTM, 140 μL of the sample was used directly for viral RNA extraction.

### Detection of SARS-CoV-2 RNA by polymerase chain reaction

SARS-CoV-2 RNA was detected by using a conventional reverse transcription polymerase chain reaction (RT-PCR) protocol adopted from the method described by Woo et al (2005) (20). The oligonucleotides used in the first round of the revised nested RT-PCR protocol targeted the same genomic region of the RNA dependent RNA polymerase (RdRp) gene described by Woo et al (2005), but were modified to be specific to SARS-CoV-2. The oligonucleotides used in the second round of PCR were newly designed. The details of the oligonucleotides are given in Table 3.

**Table 3.**
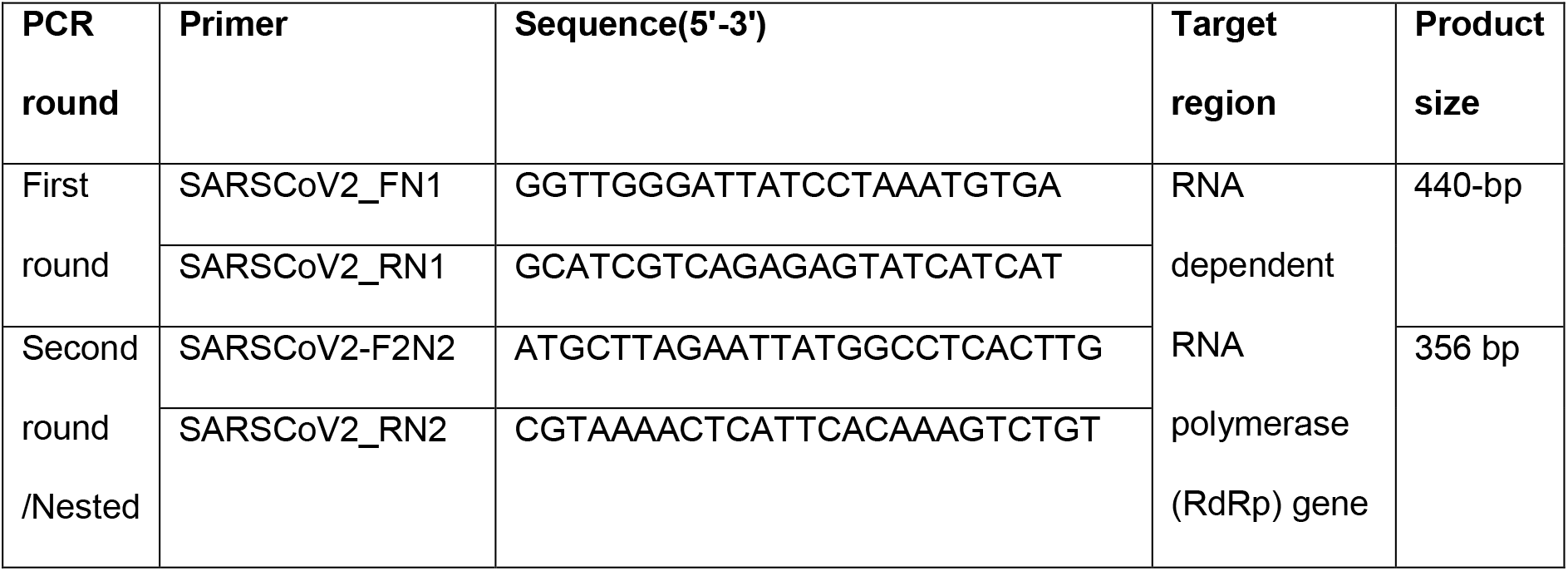
Oligonucleotides used in RT-PCR assays.

Complementary DNA (cDNA) was synthesised from extracted RNA using the Maxima H Minus First Strand cDNA Synthesis Kit (Thermo Fisher Scientific, Waltham, MA, USA) according to the manufacturer’s instructions. Targeted regions of the first round (440 bp) and second round (356 bp) of PCR were amplified using 0.5 μM of each primer pair (Table 3), 2 μl of the template and 1X Phusion™ Flash High-Fidelity PCR Master Mix (Thermo Fisher Scientific, Waltham, MA, USA). The amplification protocol for both rounds was as follows: initial denaturation at 98°C for 10 sec, 35 cycles of denaturation at 98°C for 5 sec, annealing at 61°C for 10 sec, extension at 72°C for 15 sec and final extension at 72°C for 1 min. The amplified products were visualized in 1.5% agarose gels and were purified by using Expin PCR SV mini kit (GeneAll Biotechnology, Korea) according to manufacturer’s instructions. The purified PCR products were sequenced with the respective amplicon primers at a commercial facility using the Sanger sequencing according to the BigDye Terminator Cycle Sequencing kit (Applied Biosystems, Foster City, CA, USA) protocol.

### Assembly and analysis of genome sequences

Raw nucleotide sequences were assembled using the Lasergene package version 15.0 (DNASTAR Inc., Madison, WI, USA). Contiguous sequences were aligned using BioEdit 7.0.5 software suite (21) and were used in the phylogenetic analysis to compare with global sequences retrieved from the GenBank database. The maximum likelihood tree was constructed by using the general time reversible substitution model with gamma and invariant site parameters (GTR + Gamma 5 + I), implemented in MEGA version 7 (22). The robustness of the tree was tested by bootstrapping 1000 iterations.

### Data availability

Sequences of SARS-CoV-2 generated in the present study were deposited in Genbank database under the accession numbers MT309050 and MT309051.

## Data Availability

The data or references to data supporting the findings of this study are available within the article and its supplementary materials.

## Acknowledgements

We would like to thank Dr Tan Sze Tat for his comments and suggestions on the manuscript, as well as Dr Lee Pei Xuan, Ms Jane Griffiths, Ms Praveena Jayarajah, Ms Swee Ling Low and Ms Wei Xian Luo for their support in laboratory procedures. We appreciate the Department of Public Cleanliness for their help in the coordination with premises owners. We gratefully acknowledge the authors, originating and submitting laboratories of the SARS-CoV-2 sequences in Singapore from GISAID’s EpiCoV™ Database (23) included in the phylogenetic analysis. The list of authors is detailed in Supplementary Table 2. All submitters of data may be contacted directly via the GISAID website www.gisaid.org.

## Competing interests

The authors declare no competing interests.

## Study/ethics approval

This study was approved by the Environmental Health Institute’s Management Committee (Project TS264), National Environment Agency.

## Notes

### Competing Interest Statement

The authors have declared no competing interest.

### Funding Statement

This study is internally funded.

